# Patients with affective disorders profit most from telemedical treatment: Evidence from a naturalistic patient cohort during the COVID-19 pandemic

**DOI:** 10.1101/2022.06.23.22276832

**Authors:** Tobias Rohrmann, Peter Praus, Tanja Proctor, Anastasia Benedyk, Heike Tost, Oliver Hennig, Andreas-Meyer-Lindenberg, Anna-Sophia Wahl

## Abstract

**Background:** During the COVID-19 pandemic telemedicine became essential in maintaining diagnostic procedures and treatment in psychiatry. However, it is still an open question if telemedicine is a feasible treatment option for all groups of psychiatric patients alike. This prospective monocentric observational trial was conducted to assess the general applicability of telemedical treatment in a naturalistic psychiatric outpatient cohort and to identify groups of disorders and clusters of psychopathology that respond particularly well to telemedical treatment considering sociodemographic characteristics and patients’ perspectives.

**Methods:** Patients were recruited April 2020 - April 2021 and asked to fill out the WHO-5 and the SCL-90R at baseline, after 4-6 and 8-12 weeks and a feedback-survey. Additionally, medical records, psychopathology, psychosocial functioning and sociodemographic data were analyzed. Primary outcomes were well-being, psychopathology and functioning during treatment. Secondly, diagnostic groups and psychopathology linked to a superior treatment-response were determined with respect to patients’ subjective experiences.

**Results:** Out of 1.385 patients, 254 - mostly with hyperkinetic (35.3%) and depressive disorders (24.6%) - took part. Well-being and SCL-90R total scores improved substantially (both p<0.001). CGI and GAF scores were worse in depressed subjects (both p<0.05). Improvement was mainly seen in depressed patients; chronic disorders experienced a decline in well-being. Sociodemographic characteristics could not explain this difference. Particularly female (r=0.413) patients found telepsychiatry equivalent to conventional treatment. The more virtual sessions participants attended the more likely they were to find telepsychiatry equal to conventional treatment (r=0.231).

**Conclusions:** Telemedicine is an effective treatment for patients with depression under naturalistic conditions. Telemedical consultations are a simple and reliable way of monitoring symptom severity and directing treatment choices during the treatment of depressive disorders. Patients with depression benefited more from telemedical treatment compared to participants with chronic non episodic psychiatric disorders. Future research needs to concentrate on improving telemedical treatment options suited for the latter conditions. Psychiatric telemedicine yielded overall high degrees of satisfaction among users.

## Introduction

Already at early stages of the coronavirus disease 19 (COVID-19) pandemic it became evident that an accelerated transition from face-to-face services to virtual interventions would automatically impose a plethora of critical questions concerning the adequacy and quality of such treatment options on healthcare providers, caregivers and patients alike(1). Telemedicine - in contrast to conventional psychiatric care - solely relies on the exclusive administration of mental health services via various technological devices and platforms without face-to-face contact(2). Pre-pandemic investigations had substantially focused on the applicability and potential benefits of telemedical mental health services in the context of equalizing access to these services under conditions of shortage, mainly targeting symptoms of depressive or anxiety disorders(3,4). The rapid expansion of telemedical interventions related to the COVID-19 pandemic raised the question of their comparability with conventional treatment options, hitherto mainly relying on personal relationships in the field of psychiatry and psychotherapy. Until 2020, the available data showed a steadily growing dissemination of telemedical practice, albeit only accounting for a relatively small proportion of the entirety of medical services in the U.S.(5). With the pandemic surge of COVID-19 infections in 2020, sudden changes in legislation and reimbursement practices allowed for an unprecedented expansion of telemedical treatment services in the field of psychiatry(6,7).

Besides of general advantages of telemedical treatment such as greater geographical and temporal flexibility, the reduction of age-, gender-and ethnicity-specific treatment barriers(8) and cost effectiveness(9), telemedical interventions were an essential tool in maintaining the continuity of community-based treatment paths for patients with severe mental health conditions like schizophrenia-spectrum disorders during the COVID-19 pandemic(10). Telemedical interventions also promoted the involvement of peers and relatives at multiple stages of diagnostic and treatment procedures(11).

Despite these advantages, there are some valid concerns about possible tool-related limitations of the reliability and reproducibility of telemedical assessments and categorical diagnostics under the conditions of telemedical consultations compared to traditional face-to-face evaluations(12). In particular, there is only limited evidence on the question which mental health conditions might respond optimally to exclusive telemedical treatment and which mental disorders require face-to-face appointments, respecting patients’ personal preferences. On a larger scale, telemedical interventions alone might not be suited to mitigate or overcome gross disparities in health status and access to mental health services, rather calling for thorough systematic implementation of telemedical services without marginalizing patient groups with less developed digital literacy(13).

In this longitudinal observational study, the authors aimed to elucidate the applicability, reliability and safety of telemedical assessment and treatment in a naturalistic sample of psychiatric outpatients during the COVID-19 pandemic. In this line, the purpose of this study was to identify a cluster of mental disorders and patient related features (e.g. sociodemographic characteristics) that potentially indicate a favorable response to telemedical treatment under these conditions and to determine the comparability of telemedical interventions to conventional treatments from a patient-perspective.

## Methods

### Study design

The study was designed during the first enforced Germany-wide lockdown (starting from March 22 2020) due to the COVID-19 pandemic. Most services of the outpatient clinic at the Central Institute of Mental Health, Mannheim (CIMH) at the University of Heidelberg had to be transformed into telemedical treatment options in order to maintain provision of psychiatric consultations for patients with mental health issues despite severe contact restrictions. The goal of the study was (1) to observe how psychiatric symptoms of patients with mental health problems develop during the course of telepsychiatry; (2) if patient cohorts could be identified which benefit more or less from telemedical psychiatric treatment, (3) if sex-, age or sociodemographic factors with an impact on the effectiveness of telepsychiatric treatment could be revealed and (4), how patients experienced telepsychiatric consultations compared to conventional treatment face to face with professional mental health experts.

Participants were recruited between April 2020 and April 2021. Patients received information about the study during the scheduling of their first telemedical psychiatric counseling. The aims and purpose of the study were explained either by members of the study team who contacted interested patients or by the psychiatrist or psychologist who provided the first telemedical session. Patients who gave informed consent to participate were asked to complete three surveys during the course of the study: Before the start of the first telemedical consultation participants agreed that their medical record which would be created during telemedical treatment could be used for further analysis (see below) as part of the study. Subjects were also asked to fill out the WHO-5 well-being index (WHO-5(14)) and the symptom check-list-90-R (SCL-90R(15)), which were provided paper-based. 4-6 and 8-12 weeks after the first telemedical counseling session, participants could choose for the second and third survey, if they preferred an interview via phone call or online surveys using REDCap (https://www.project-redcap.org/), a secure web application for building and managing online surveys for research studies and operations supported by the National Institutes of Health (NIH/NCATS UL1 TR000445). N=254 participants returned the first paper-based survey (including 9 anonymous study subjects). N=94 participated in the second survey via RedCap and N=50 via telephone. N=68 study subjects also completed the third survey via RedCap, while N=48 were interviewed via telephone. During the second and third survey patients were asked to fill out the WHO-5 questionnaire again while participants were only requested twice to complete the SCL90-R (1^st^ and 2^nd^ survey).Either during the second or the third survey patients could evaluate the telemedical psychiatric counseling (see details below).

### Acquisition psychiatric history and sociodemographic data

N=182 subjects (71.7%) gave informed consent to acquire sex, age and sociodemographic data as well as psychiatric history and psychopathological symptoms from the medical record. All professional experts providing telepsychiatric services in our outpatient clinic were instructed to use a structured form in the computerized documentation system of our clinic (ORBIS, SAP, Walldorf Germany) for the first interview. Patients were asked for current psychiatric symptoms, psychiatric diagnoses according to the ICD-classification of the WHO (version 10), past psychiatric history and sociodemographic data such as current life and living situation, education, professional training and labor situation, debts and history of criminal assaults. All psychiatrists and psychologists of our outpatient clinic were also requested to score patients on initial appearance according to the global assessment of functioning scale (GAF(16)) and the clinical global impression scale (CGI(17)). For later data analysis electronic medical records were systematically queried for the number of telemedical treatment sessions participants received and possible hospitalizations during the course of the study. Medical records were additionally scrutinized for the number of in- and outpatient treatments and days of hospitalization during the year before March 2020, when outpatient psychiatric care was still provided personally.

### Evaluation and follow-up of psychopathological symptoms

#### WHO-5 Well-Being Index (WHO-5)

For assessment of overall well-being over the course of telepsychiatric counseling the WHO-5, a short self-administered measure of well-being over the last two weeks(18), was used. The WHO-5 consists of five positively worded items that are rated on a 6-point Likert scale, ranging from 0 (at no the time) to 5 (all of the time). We transformed the raw scores to a score from 0 to 100 (raw data*4), where lower scores indicated worse well-being. A score of ≤50 was considered as poor wellbeing and a score of 28 or below indicative of depression.

### SCL-90-R

The SCL-90-R by Derogatis(19) measures the subjective perception of physical and mental symptoms a person has experienced during the past seven days. All 90 symptoms are scored on a Likert scale consisting of 5 steps, ranging from 0 (the symptom is not there at all) to 4 (indicating very strong impairment due to the symptom). We analyzed the data gained in two surveys of the SCL-90-R according to the instructions provided by Derogatis/Franke, German Version, 2^nd^ Edition, Beltz Test, 2000. T-Values equal to and above 60 were considered as a relevant mental detraction from the respective symptom or global score.

### Clinical Global Impression Scale (CGI) and the global assessment of functioning scale (GAF)

While the WHO-5 and the SCL-90-R are self-report questionnaires on the subjective perception of overall well-being and different symptom domains, the GAF and CGI were used as clinician-rated scales to document the global impairment due to patients’ (mental) health conditions. The CGI scores the severity of the symptoms, ranging from 1 to 7 (1= normal/not at affected; 7=very severely ill(17)). The GAF indicates the global functioning of a patient taking into account the psychiatric, social and professional level. The scale is ranging from 0 (very sick) to 100 (healthy)(16).

### Evaluation of telemedicine by participants

N=112 (44.1% of all participants) completed an evaluation questionnaire asking for feedback concerning technical details (e.g. if patients decided for phone call or video conference or both and if interruptions occurred due to technical problems) and the overall experience with the telepsychiatric consultations. Participants were asked how helpful they found the telemedical intervention and if it was comparable to a conventional face-to face contact. Patients were also requested to state their preference about using telepsychiatry in the future again. Participants could also document pros and cons of the telemedicine and their wishes for future improvements.

### Data analysis

All data acquired during surveys and from the medical records were entered into an Excel master file and then translated to SPSS Version 27 and R Version 4.1.1. for further analysis. The descriptive statistics of the sample were computed for the sociodemographic characteristics, consisting of frequencies and percentages for categorical values and mean and standard deviations (SD) for scale variables. Differences between two groups of patients (depressive and suffering from other psychiatric diseases) were assessed using the Mann-Witney test for the different non-parametric clinical scale variables. The differences in these scales over time for the single patients were assessed with an ANOVA test and a post-hoc Bonferroni test.

A paired Wilcoxon signed-rank test was used to assess differences in the levels of mental health variables such as e.g.: WHO-5 or SCL 90 comparing the results of the three inquiries. Bivariate associations between WHO-5 and SCL90-R, CGI and GAF were analyzed using linear regression models and between mental x and y (continuous variable) were assessed via Spearman’s correlation coefficient *r*. For all tests, a p-value of less than 0.05 was considered statistically significant.

### Ethics Approval and Consent to Participate

Written informed consent was obtained from all subjects who participated in our study, except for 9 cases where patients sent the paper-based WHO-5 and SCL90-R questionnaires of the first survey back to us anonymously. These data could not be further evaluated. The study design and data acquisition were presented to the ethics committee II at the medical faculty Mannheim, University of Heidelberg and approved (No. 2020-562N).

## Results

During the first year of the Covid-19 pandemic (April 2020 - April 2021) 8.235 telemedical treatment sessions were provided to 1.385 patients via phone or video call by the psychiatrists and psychologists of the outpatient clinic at the Central Institute of Mental Health (CIMH) in Mannheim, Germany. 254 (18.3%) of these patients gave their informed consent to participate in our study and to use their medical record generated during telemedical treatment for further systematic analysis. Out of 239 patients who engaged in the first survey at baseline, 140 patients responded to the second, and 103 patients to the third survey 4-6 and 8-12 weeks after the first telemedicine session, respectively.

### Demographics and psychiatric history

In total, electronic medical records of 174 patients could be evaluated revealing sociodemographic characteristics as presented in Table 1: Our study population consisted of younger adults, 38.6 ± 13.1 years old with more women (56.7%, 39.9 ± 13.3 years old) than men (37,8%, 37.0 ± 12.6 years old) participating. Most patients had a partner (55.3%), but no children (82.8%) and were living alone (53.6%). Almost half of the study population (48.8%) was well educated with at least 12 years of school education while more than half (53.8%) had also successfully completed apprenticeship. 53.3% were employed during the time of the telemedicial counseling and most patients (63.9%) lived in a stable financial situation without significant debts.

**Table 1.**
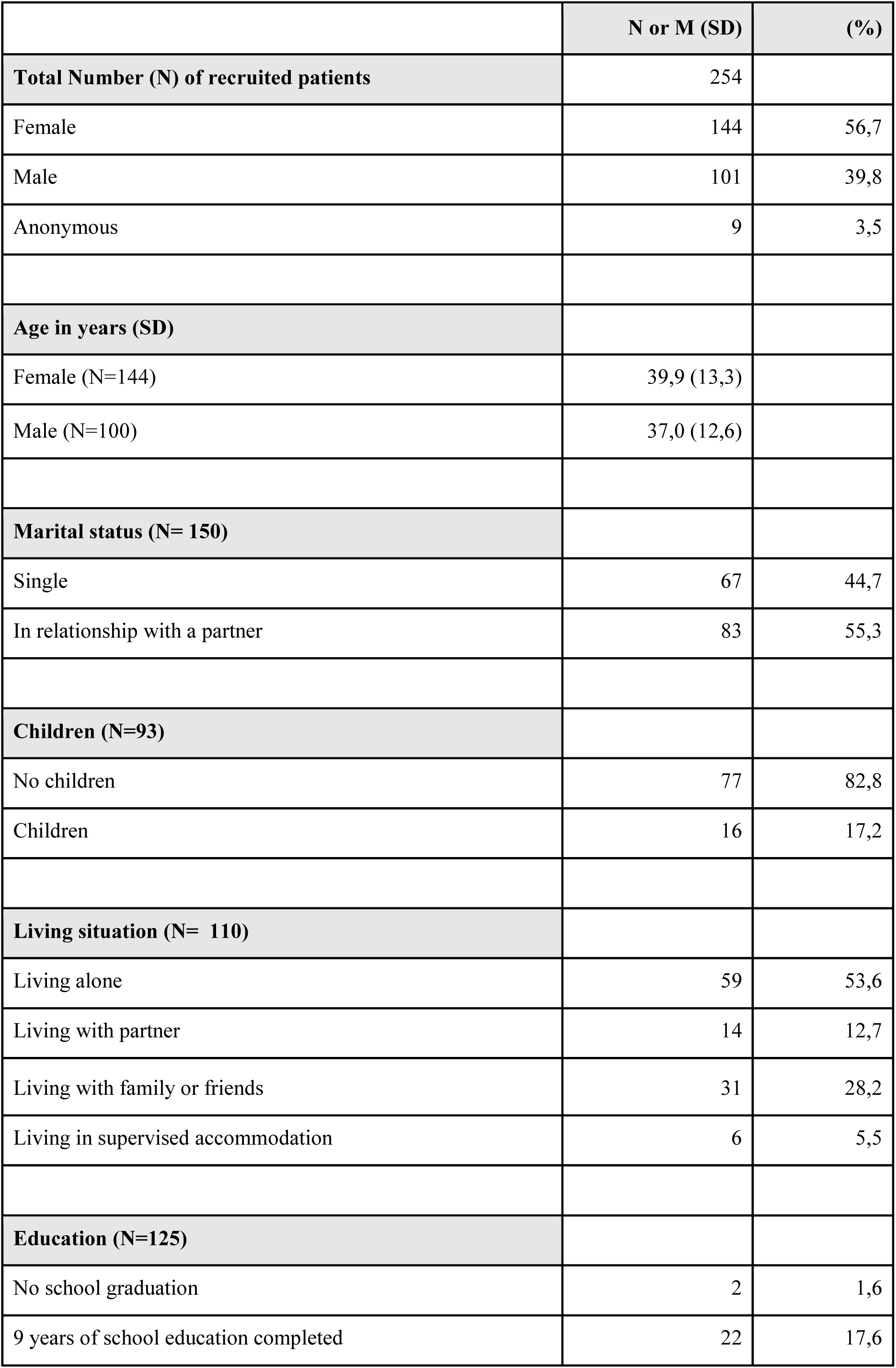
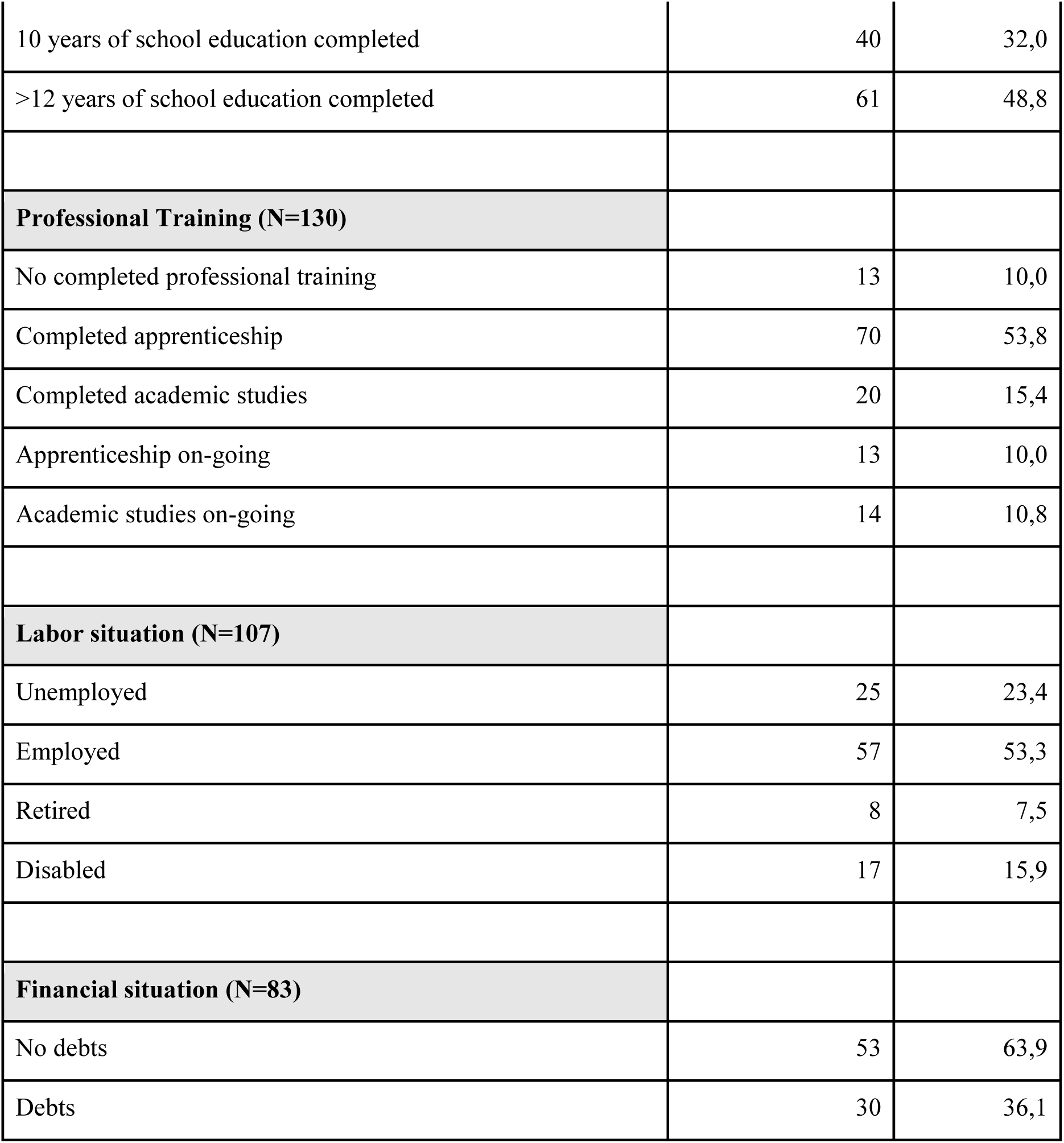
Sociodemographic characteristics of the study population assessing sex, age, marital status, children, living situation, eduction, professional training, labor and financial situation. N= number of subjects where information was found in the medical record. The percentage was calculated as (N/N responded)*100. M=mean, SD= standard deviation.

|  | N or M (SD) | (%) |
| --- | --- | --- |
| **Total Number (N) of recruited patients** | 254 |  |
| Female | 144 | 56,7 |
| Male | 101 | 39,8 |
| Anonymous | 9 | 3,5 |
| **Age in years (SD)** |  |  |
| Female (N=144) | 39,9 (13,3) |  |
| Male (N=100) | 37,0 (12,6) |  |
| **Marital status (N= 150)** |  |  |
| Single | 67 | 44,7 |
| In relationship with a partner | 83 | 55,3 |
| **Children (N=93)** |  |  |
| No children | 77 | 82,8 |
| Children | 16 | 17,2 |
| **Living situation (N= 110)** |  |  |
| Living alone | 59 | 53,6 |
| Living with partner | 14 | 12,7 |
| Living with family or friends | 31 | 28,2 |
| Living in supervised accommodation | 6 | 5,5 |
| **Education (N=125)** |  |  |
| No school graduation | 2 | 1,6 |
| 9 years of school education completed | 22 | 17,6 |
| 10 years of school education completed | 40 | 32,0 |
| >12 years of school education completed | 61 | 48,8 |
| **Professional Training (N=130)** |  |  |
| No completed professional training | 13 | 10,0 |
| Completed apprenticeship | 70 | 53,8 |
| Completed academic studies | 20 | 15,4 |
| Apprenticeship on-going | 13 | 10,0 |
| Academic studies on-going | 14 | 10,8 |
| **Labor situation (N=107)** |  |  |
| Unemployed | 25 | 23,4 |
| Employed | 57 | 53,3 |
| Retired | 8 | 7,5 |
| Disabled | 17 | 15,9 |
| **Financial situation (N=83)** |  |  |
| No debts | 53 | 63,9 |
| Debts | 30 | 36,1 |

All patients included in the study were diagnosed with at least one psychiatric disorder (Table 2): The largest group (35.3%) consisted of patients with hyperkinetic and tic disorders, while almost a quarter of the study population (24.6%) was primarily diagnosed with a depressive episode or recurrent depressive disorder according to ICD-10. In addition, for 32 patients (12,6%) a depressive or recurrent depressive disorder was noted as secondary diagnosis. Schizophrenia was only diagnosed in 3 cases (1.3%) as a main psychiatric disorder justifying current psychiatric treatment. Matching with this preponderance of affective and neurodevelopmental disorders, most patients presented with psychopathological symptoms (Supplementary Table 1) typical of affective and hyperkinetic disorders such as attention (51.9%) and concentration deficits (67.6%), abnormal thought processes (71.7%), changes in mood (84.0%), reduced ability to experience joy (52.9%), lack of drive (56.9%), anxiety (64.4%) and sleep disturbances (69.4%). Orientation, mnestic functioning as well as illness insight were undisturbed in almost all patients (see Supplementary Table 1), while symptoms typical of an episode of psychosis such as abnormal thought content, hallucinations or intrusions were almost absent (Supplementary Table 1). Among the patients who gave full particulars of their psychiatric medical record (28.3%), 29.2% reported a history of attempted suicides. Three quarters of participants (74.3%) had been already treated for psychiatric issues in the past, while only a minority (25.8%) received psychiatric treatment for the first time.

**Table 2.**
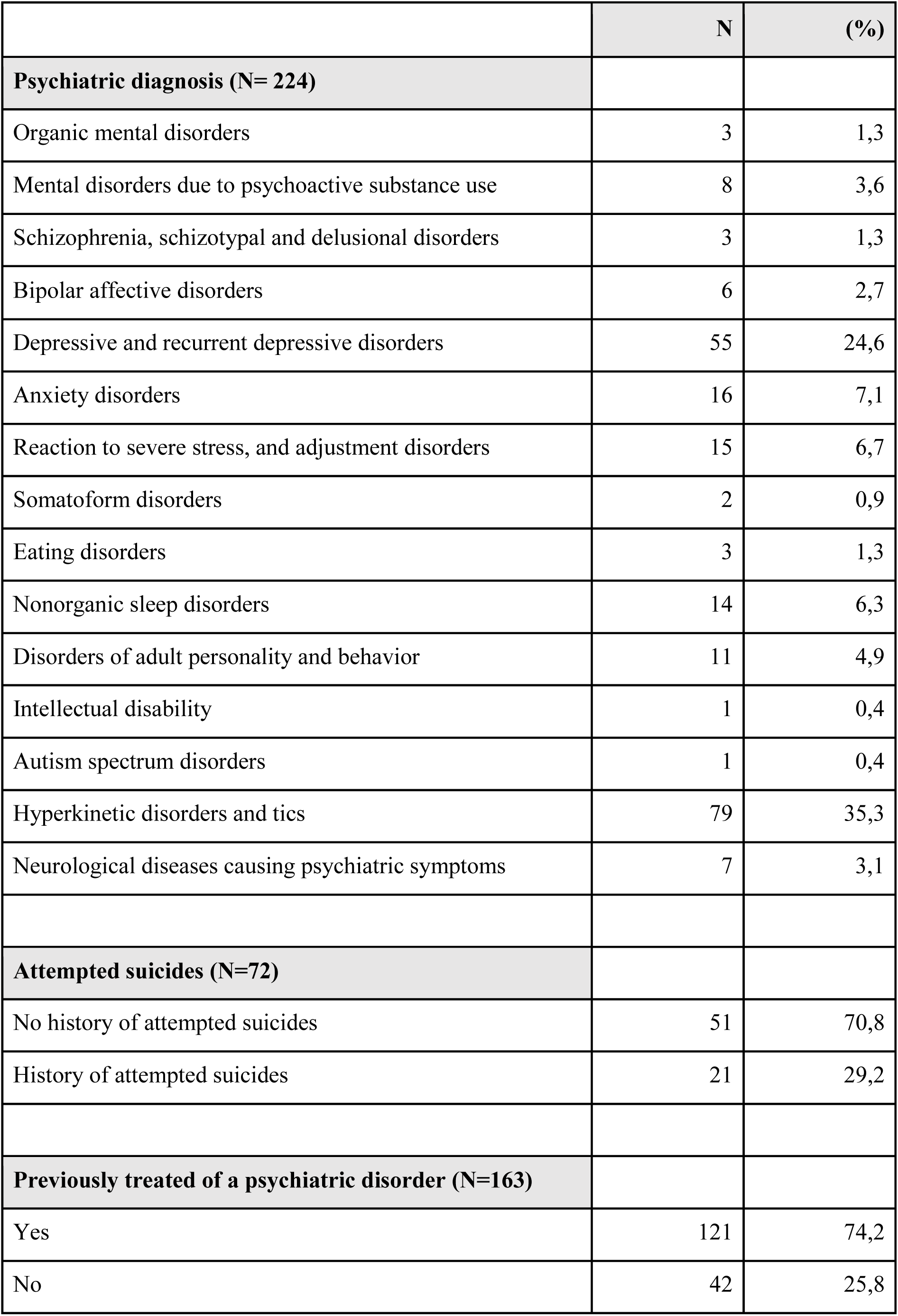
Table depicting the psychiatric history of subjects including the leading psychiatric diagnosis documented in the medical record, as well as attempts of suicides and previous psychiatric treatments. N= number of subjects where information was found in the medical record. The percentage was calculated as (N/N responded)*100.

|  | N | (%) |
| --- | --- | --- |
| **Psychiatric diagnosis (N= 224)** |  |  |
| Organic mental disorders | 3 | 1,3 |
| Mental disorders due to psychoactive substance use | 8 | 3,6 |
| Schizophrenia, schizotypal and delusional disorders | 3 | 1,3 |
| Bipolar affective disorders | 6 | 2,7 |
| Depressive and recurrent depressive disorders | 55 | 24,6 |
| Anxiety disorders | 16 | 7,1 |
| Reaction to severe stress, and adjustment disorders | 15 | 6,7 |
| Somatoform disorders | 2 | 0,9 |
| Eating disorders | 3 | 1,3 |
| Nonorganic sleep disorders | 14 | 6,3 |
| Disorders of adult personality and behavior | 11 | 4,9 |
| Intellectual disability | 1 | 0,4 |
| Autism spectrum disorders | 1 | 0,4 |
| Hyperkinetic disorders and tics | 79 | 35,3 |
| Neurological diseases causing psychiatric symptoms | 7 | 3,1 |
| **Attempted suicides (N=72)** |  |  |
| No history of attempted suicides | 51 | 70,8 |
| History of attempted suicides | 21 | 29,2 |
| **Previously treated of a psychiatric disorder (N=163)** |  |  |
| Yes | 121 | 74,2 |
| No | 42 | 25,8 |

Most patients (93,7%, Table 3) had not been in contact with our outpatient clinic within 12 months before the pandemic. During the course of the study patients had on average 4 (4.26 ± 5.45) psychiatric appointments. At least 3 (3.05 ± 3.58) appointments were provided via phone or video call. For comparison, the 16 patients that had already been in psychiatric outpatient treatment before the pandemic had also had 4 (4.56 ± 5.89) face to face appointments during the course of 12 months before the pandemic. Only a small number of participants (n=13, 5.12%) required hospitalization during the study period. These 13 patients spent on average 74.54 ± 46.67 days on a psychiatric ward (Table 3), while four participants had stayed in hospital more than 4 months (128 ± 62.20) in the year prior to the pandemic.

**Table 3.** Table revealing the amount of psychiatric counseling, hospitalization requirements and days of hospitalization in a specialized psychiatric unit 12 months before the pandemic and during the course of the study in the Covid-19 pandemic. N= number of subjects where information was found in the medical record. The percentage was calculated as (N/N responded)*100. M=mean, SD= standard deviation.

| Out-patient treatment (Mean # of doctoral appointments) | M (SD) |
| --- | --- |
| Within 12 months before the pandemic (N=16) | 4,56 (5,89) |
| During the pandemic (N=254) | 4,26 (5,45) |
| via telemedicine | 3,05 (3,58) |
| with personal contact | 1,25 (2,98) |
| Patients requiring in-patient treatment (N=254) | N (%) |
| Within 12 months before the pandemic | 4 (1,57) |
| During the pandemic | 13 (5,12) |
| Mean - days of hospitalization (N=254) | M (SD) |
| Within 12 months before the pandemic (N=4) | 128 (62,20) |
| During the pandemic (N=13) | 74,54 (46,67) |

### Changes of psychopathology according to WHO-5 and SCL90 self-assessment under telemedicine psychiatry

Subjects were requested to report current mental well-being by completing the WHO-5 Well-Being Index (WHO-5,(18)) before, as well as 4-6 and 8-12 weeks after the beginning of telepsychiatric treatment. 237 patients (93.3%) participated in the first self-report, scoring 31.65 ± 20.11 (range 0-100, Fig. 1A). The prevalence of poor well-being (WHO-5 score≤50, (20)) was 81.9% (n=194) and that of depression (WHO-5 score≤28) was 55.3% (n=131) before the start of the telemedical treatment. 131 (51.6%) and 98 (38.5%) of the initial 254 participants also responded to the second and third survey, respectively. Patients reported a significant improvement of well-being 4-6 and in particular 8-12 weeks after the start of the telemedical appointments (WHO-5 4-6 weeks 39.60 ± 21.61; WHO-5 8-12 weeks 41.47 ± 21.26; p < 0.001, the Wilcoxon test was used for comparison with the initial WHO-5 self-report). 8-12 weeks after initiation of the telemedical treatment only 32% (n = 33) revealed very poor well-being, indicative of severe depressive symptoms (WHO-5 score≤28).

**Figure 1.**
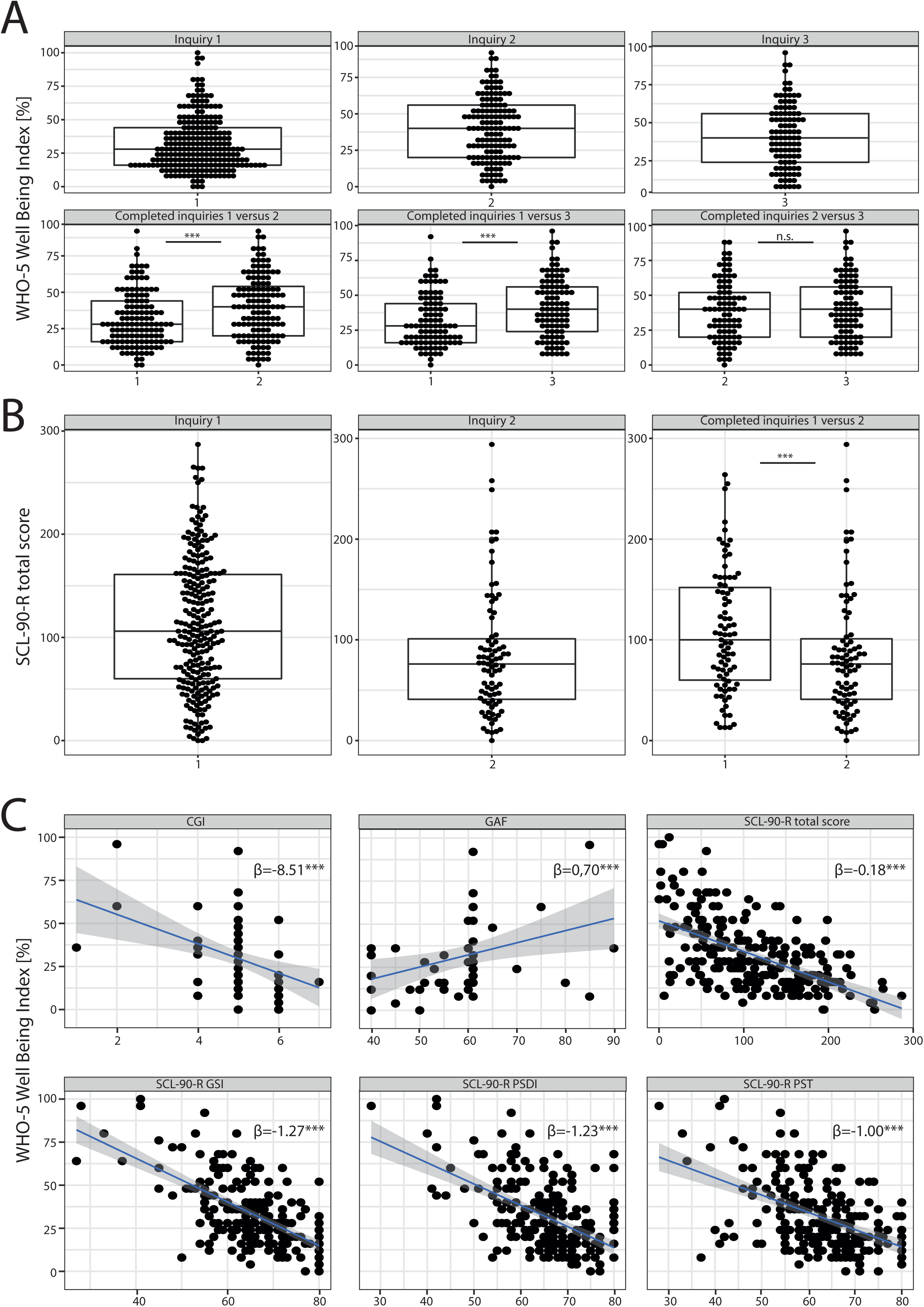
Results of self-report assessment using WHO-5 and SCL90-R. (A) Results of the WHO-5 Wellbeing Index: Boxplots with scoring details of all individuals participating in the different surveys (inquiry 1= survey before the beginning of the telemedical treatment; inquiry 2 and 3= surveys 4-6 and 8-12 weeks after start of the telemedical counseling respectively) are shown in the upper panel. In the lower panel we compared the results of subjects who participated at least in the first and one consecutive survey. We found a significant improvement of WHO5 scores during the course of telemedical treatment (inquiry 1 versus 2, p<0.001, Wilcoxon-test). (B) Results of the SCL90-R. Those participants that completed the survey twice (before and 4-6 weeks after start of the telemedical psychiatry) reported a significant improvement of psychopathological symptoms (significant decrease of total scores for inquiry 2 versus 1, p<0.001, Wilcoxon-test). (C) Results of the WHO-5 correlated to clinical scores such as the CGI and the GAF as well as to all three global indices of distress of the SCL90-R such as the GSI, PSDI and PST (A linear regression analysis was performed, β=regression coefficient). ***p<0.001. n.s.= non-significant.

Patients were also asked to fill out the Symptom Checklist-90 Revised (SCL90-R) at baseline, and 4-6 weeks after the start of the telemedical treatment. The mean SCL90 total score for n = 83 (32.68%) participants was 107.9 ± 60.86 at baseline (1^st^ inquiry, Fig. 1B) with a decline to a mean score of 85.08 ± 61.22 in the second survey (2nd inquiry, Fig. 1B), indicating a significant improvement of psychopathological symptoms after 4-6 weeks (p<0.001 with a paired Wilcoxon test to compare the SCL90 total scores of patients who participated in both inquiries). For patients who participated in both inquiries (n = 86), the means of all three global indices of distress in the first inquiry were above the critical threshold (T-value **≥** 60, Supplementary Figure 2). The Global Severity Index (GSI) was 66.61 ± 10.31. 77% (n = 83) of the study population who participated in both inquiries scored with a T-value **≥** 60 suggesting “psychologically measurable distressed cases” (21). In addition, the Positive Symptom Distress Index (PSDI) and the Positive Symptom Total (PST) were evaluated as global measures of symptom intensity and number of psychiatric symptoms with significant burden. Means of both indices revealed T-values **≥** 60 (PSDI 65.12 ± 8.23; PST 62.02 ± 8.45) substantiating significant mental distress. Furthermore, all mean values of the nine primary symptom dimensions (Supplementary Figure 2) exceeded T-values of 60. In particular the subscales for obsessive-compulsive symptoms and depression scored even above a T-value **≥** 65 (Mean obsessive-compulsive 67.55 ± 10.77; Mean depression 67.08 ± 10.61, Supplementary Figure 1), revealing significant burden in the study population (T ≥ 1½ SD of mean T-values in a healthy norm group, Franke 2001). All global scales and nine primary dimension scores were markedly decreased in the second inquiry (p<0.001); a Wilcoxon test was used for the comparison of the results of patients with both inquiries completed (Supplementary Figure 2). In particular T-Values were below 60 for the PSDI (59.45± 10.67) and for 5 out of nine primary dimension scores (anxiety, hostility, phobic anxiety, paranoid ideation and psychoticism, Supplementary Fig. 2), indicating a reduction of psychopathological burden for these dimensions to the range of a healthy norm group.

The correlations of the WHO-5 self-assessments with the SCL90-R global scores are displayed in Fig. 1C. With regard to well-established clinical scales, relationships between the response and explanatory variables were obtained; for the Clinical Global Impressions Scale (CGI), WHO-5 decreased by 8.51 per CGI increase to measure the severity of psychopathology and for the Global Assessment of Functioning scale (GAF), the WHO-5 increased by 0.70 points per GAF point increase.

### Patients diagnosed with depression profit more from telemedicine psychiatry than those with other psychiatric entities

We next investigated how patients diagnosed with a depressive episode developed under telepsychiatric treatment, as these patients were significantly more severely affected based on clinical judgment than patients with other psychiatric entities: The CGI was significantly higher for patients with depression (4.98 ± 0.89) than for those without (4.70 ± 0.92, p<0.05, Mann-Whitney test, Fig. 2A). In addition, the GAF was significantly lower for depressive subjects (57.46 ± 10.77) than for those participants with other psychiatric issues (62.18 ± 10.77, p<0.05, Mann-Whitney test, Fig. 2B). Depressive patients also scored significantly lower in the WHO-5 in the first inquiry before the start of the telemedical treatment (Fig. 2C, depressive patients 18.80±14.57 versus others 30.92 ± 22.00, p<0.001, two-way repeated measures analysis of variance (ANOVA) with post hoc Bonferroni). However, during the course of telepsychiatric treatment the WHO-5 improved for depressed patients to a level comparable to patients with other psychiatric disorders, who were clinically less severely ill at the beginning of treatment (Fig. 2C, p=0.22 for inquiry 2 comparing patients with and without depression and p>0.99 for inquiry 3, ANOVA with post hoc Bonferroni). In particular, patients with depression reported an improvement of >20% on the WHO-5, while subjects with chronic disorders like personality disorders or hyperkinetic disorders who completed the 2nd and 3rd survey of the WHO-5 indicated a decline in mental well-being (Fig. 2D). We also investigated potentially protective or supportive sociodemographic characteristics of depressed patients that were lacking in other patients. However, when examining life and labor situation, education and professional training, children and living situation, as well as debts and the history of criminal assaults, we did not find significant differences (Fig. 2E).

**Figure 2.**
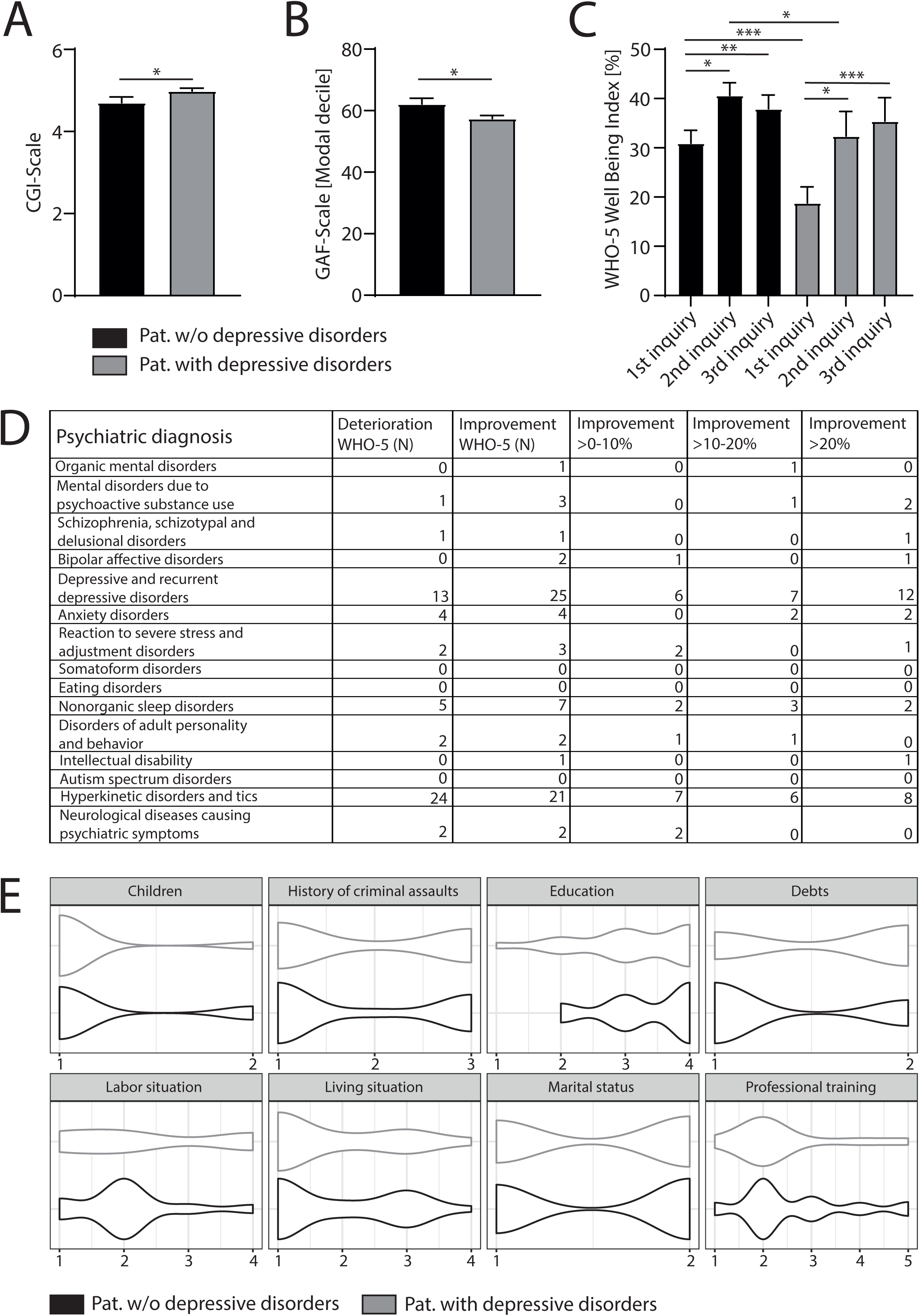
Based on clinical judgment using the clinical scores, CGI (A) and GAF (B) patients diagnosed with depression were significantly sicker than subjects with other psychiatric entities (for both CGI and GAF Mann-Whitney test was used to compare participants with and without a diagnosis of depression). (C) Although depressive patients scored lower in the WHO-5 than patients with other psychiatric entities (results inquiry 1 of depressive patients versus others, p<0.001, ANOVA with post hoc Bonferroni), overall well-being of this patient cohort improved in such a way, that during the 2nd and 3rd inquiry, WHO-5 total scores significantly raised (inquiry 1 versus 2, p<0.05; inquiry 1 versus 3, p<0.001, ANOVA with post hoc Bonferroni) to the same levels as found in patients without a diagnosis of depression. (D) Table depicting the number of participants diagnosed with different psychiatric entities and how many of these subjects reported a decline or improvement in well-being according to the WHO-5 during the course of the telemedical counseling. (E) Violin plots showing sociodemographic characteristics for the two cohorts (subjects diagnosed with depression and those without). Using the same different subcategories as described in table 1 we did not find a significant difference between the distribution of sociodemographic characteristics. Results are presented in mean ± SEM; *P<0.05, **P<0.01, ***P<0.001.

### Satisfaction with telemedicine psychiatry

At the end of our observational study, patients were asked to evaluate the telepsychiatric treatment provided during the study period. Most consultations were arranged via phone (89.29%, Table 4) and technical problems were rarely an issue (87.50%). More than 80% of participants (81.48%, Table 4) were satisfied with telemedical treatment. 49.07% (Table 4) of subjects experienced telepsychiatric counseling as effective as face-to-face treatment. Almost half of patients (49.54%, Table 4) considered using telemedicine in the future again. In particular female patients rated telemedical treatment as equivalent to counseling in person (r = 0.413, Spearman’s correlation) and expressed their wish to use telemedicine in the future again (r = 0.342, Spearman’s correlation). The more telepsychiatric sessions subjects attended the more likely they were to experience telepsychiatry as equal to conventional outpatient therapy (r = 0.231, Spearman’s correlation). When participants were asked to describe pros and cons of telepsychiatry, half of the participants providing feedback (N=102 statements) missed the personal atmosphere when talking face to face, while a quarter even reported to have missed substantial information of the conversations due to missing gestures and body language. 16.7% could not describe any disadvantage at all. 46.1% found telepsychiatry much more comfortable than conventional therapy, as it was secure to prevent any kind of infection (10.8%) and required less time. Patients also reported (27.4%) to feel less stressed during telepsychiatric treatment when providing intimate details of their biography while sitting in a familiar environment at home.

**Table 4.**
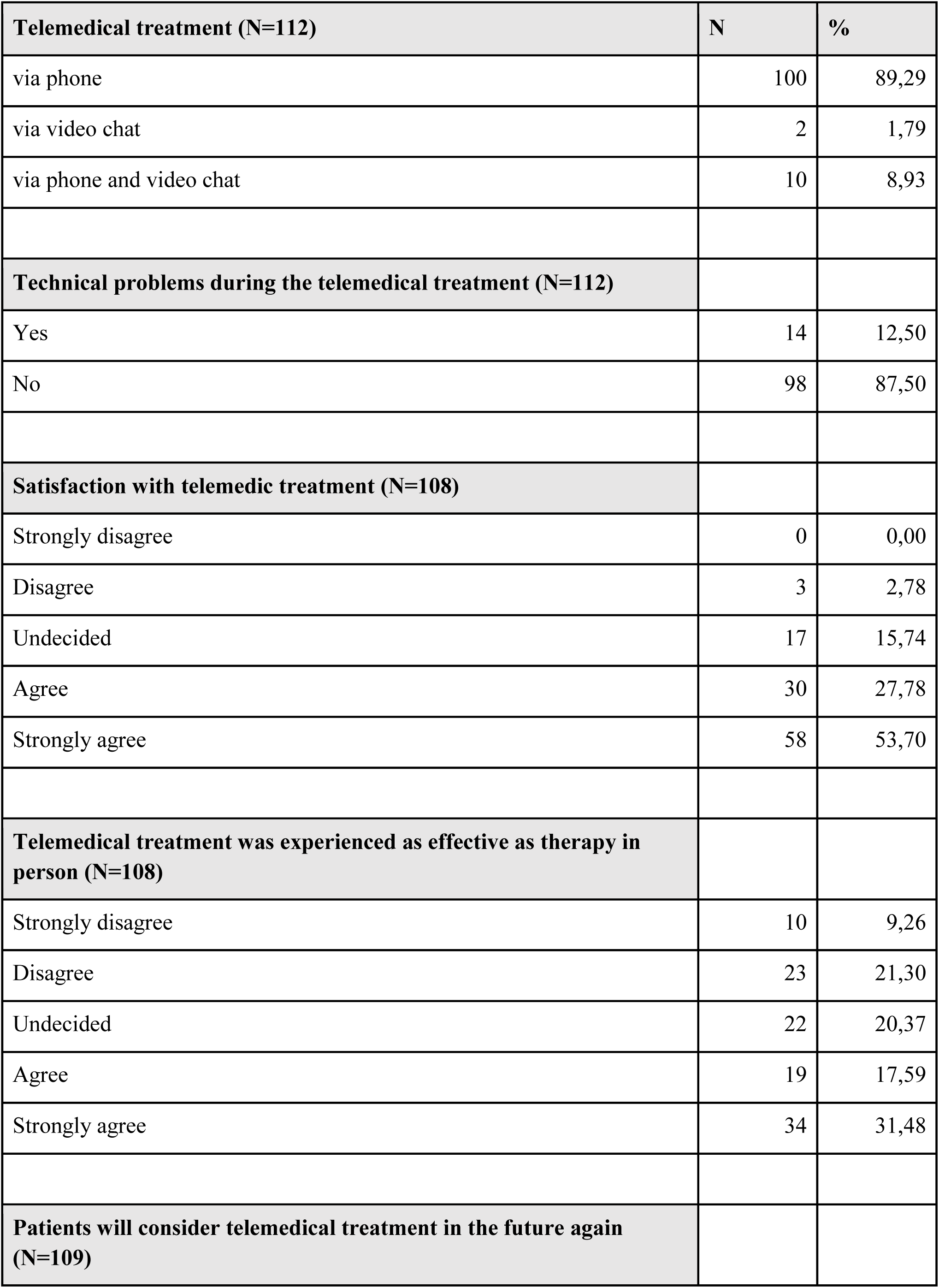
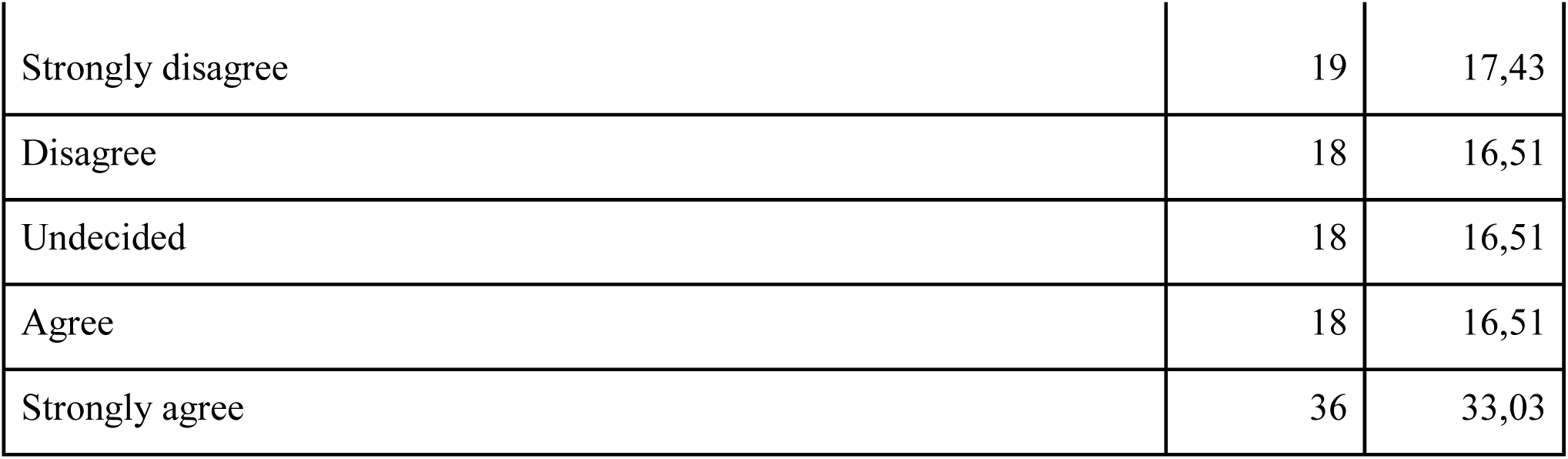
Results of the evaluation of the telemedical psychiatric counseling by the study participants requesting feedback considering the mode of telemedical treatment, problems that emerged during telemedical treatment, overall satisfaction and if subjects are willing to use telemedical offers in the future. N= number of subjects that provided feedback at the end of the telemedical treatment. The percentage was calculated as (N/N responded)*100.

| **Telemedical treatment (N=112)** | **N** | **%** |
| --- | --- | --- |
| via phone | 100 | 89,29 |
| via video chat | 2 | 1,79 |
| via phone and video chat | 10 | 8,93 |
| **Technical problems during the telemedical treatment (N=112)** |  |  |
| Yes | 14 | 12,50 |
| No | 98 | 87,50 |
| **Satisfaction with telemedic treatment (N=108)** |  |  |
| Strongly disagree | 0 | 0,00 |
| Disagree | 3 | 2,78 |
| Undecided | 17 | 15,74 |
| Agree | 30 | 27,78 |
| Strongly agree | 58 | 53,70 |
| **Telemedical treatment was experienced as effective as therapy in person (N=108)** |  |  |
| Strongly disagree | 10 | 9,26 |
| Disagree | 23 | 21,30 |
| Undecided | 22 | 20,37 |
| Agree | 19 | 17,59 |
| Strongly agree | 34 | 31,48 |
| **Patients will consider telemedical treatment in the future again (N=109)** |  |  |
| Strongly disagree | 19 | 17,43 |
| Disagree | 18 | 16,51 |
| Undecided | 18 | 16,51 |
| Agree | 18 | 16,51 |
| Strongly agree | 36 | 33,03 |

## 4 Discussion

The current study aimed at longitudinally investigating the applicability and effectiveness of telemedical treatment in a naturalistic monocentric sample of psychiatric outpatients during the first waves of the COVID-19 pandemic in Germany. A secondary aim was to identify clusters of psychopathology and determine sociodemographic features that were indicative of a favorable response to telemedical psychiatric outpatient treatment. Lastly, the study addressed patients’ perceptions of telemedical treatment.

More than one third of participants was diagnosed with a hyperkinetic disorder, mostly attention deficit hyperactivity disorder (ADHD) or attention deficit disorder (ADD), probably due to the professional specialization of general psychiatric outpatient services at the CIMH. Almost a quarter of participants was diagnosed with a depressive episode as the main diagnosis during the treatment interval of concern. However, psychopathology in the entire sample, as rated by clinicians, was dominated by symptoms of depression and anxiety in a majority of participants at the onset of telemedical treatment. This is consistent with earlier findings concerning psychological distress linked to the outbreak of the COVID-19 pandemic(22).

Most participants reported poor well-being and high rates of depression before the onset of telemedical psychiatric outpatient treatment on the WHO-5 and showed significant improvement 8-12 weeks after the initiation of telemedical treatment. Furthermore, the study population showed a significant degree of psychological distress as measured by the SCL 90-R, both on a global scale (GSI) as well as concerning symptom intensity (PSDI) and quantitative symptom load (PST). In this line, participants’ self-assessments via the SCL 90-R revealed a significant burden in all nine primary symptom dimensions of the SCL 90-R, centering around the subscales for depressive and obsessive-compulsive symptoms at baseline. After 4-6 weeks a significant reduction of psychopathology as measured by the SCL 90-R was observed, partly equivalent to the range of a healthy norm group. Generally, patients’ self-assessment via the WHO-5 and the SCL 90-R correlated well with observer-rated impressions on more generic measures of clinical functioning and disorder-severity (CGI, GAF). Thus, the specific conditions of telepsychiatric assessment and treatment did not impede the validity and reproducibility of diagnostic procedures in the study sample. This corroborates earlier findings that the implementation of patient reported outcome measures via telemedicine is a viable way of assessing psychiatric symptoms and psychosocial functioning as well as monitoring clinical changes in order to improve treatment outcomes(23).

Overall, participants diagnosed with depressive disorders during telemedical treatment were significantly more affected by a deterioration of well-being and functioning than participants suffering from other mental disorders. Yet, during the treatment process, patients with depressive disorders showed a pronounced improvement, whereas individuals with chronic or non-episodic disorders like ADHD and personality disorders did not benefit likewise and - somewhat counterintuitively - showed a decline in mental well-being on the WHO-5 during the study period. Specific sociodemographic traits derived from patients’ electronic medical records were not associated with this remarkable difference in treatment outcome, although it could be speculated that chronically impaired individuals might be exposed to more challenging social and environmental conditions (e.g. societal isolation, job insecurity, downward social mobility). Patients with ADHD and other neurodevelopmental disorders may generally be more vulnerable to a variety of negative health outcomes and increased mortality(24). ADHD symptom severity predicted adherence to preventive measures during the pandemic and higher psychological distress(25). Yet, it is unknown if the ADHD patients in our sample experienced higher or equal levels of distress and higher levels of well-being before the pandemic. Thus, variance and oscillations of psychopathology in neurodevelopmental disorders that are predominantly chronic by nature could not be effectively influenced during the study period by telemedical treatment in our sample. In contrast, available data concerning the telemedical treatment of chronic somatic diseases points at an overall positive effect of telemedicine on the management of these conditions(25).

Overall, a vast majority of participants reported high satisfaction with and robust acceptance of the telemedical treatment administered during the study period. Roughly 50% of participants, predominantly female, rated telemedical treatment equivalent to treatment provided face to face. This finding seems to be of particular interest as almost 75% of all participants had undergone psychiatric treatment at some point before their participation in this study. Remarkably, patients’ satisfaction with telemedical treatment seemed to increase with the number of telemedical consultations. This is in line with findings from a current nationwide multicentric study from Germany where overall good experiences with the telemedical treatment of affective, stress related and somatoform disorders were reported(26).

### Limitations

There are some limitations concerning the above mentioned results that warrant further discussion. Firstly, the study sample might not be fully representative of the entirety of patients that underwent telemedical treatment during the COVID-19 pandemic at the CIMH. The study sample could be enriched for individuals with an above average willingness to partake in an observational study with repeated surveys. Consequently, severely and extremely mentally ill patients who experience difficulties filling out questionnaires and checklists repeatedly might have been excluded. An earlier study found that older patients and patients with more severe disorders (e.g. schizophrenia) were less likely to use telemedicine, whereas female patients with anxiety and depressive disorders as well as post-traumatic stress disorder (PTSD) more frequently used telemedicine(27). Furthermore, participants were not screened for their cultural or ethnic background. Therefore it cannot be excluded that language or cultural barriers prevented potential participants from being included into the study cohort as described before in a sample of African American patients from North America(7). There is no information regarding the degree of digital literacy in the study sample although it is characterized by a relatively high degree of education. Previous studies demonstrated that healthcare providers’ degree of experience with digital technologies is associated with the willingness to use these technologies and the way they are judged by their users(28). Likewise, the acceptance of telemedicine by its users is shaped by the perceived usefulness of the technologies applied, social influence and personal attitude(29).

It cannot be ruled out that the significant improvement in psychopathology and functioning observed in individuals with depressive disorders during telemedical treatment merely corresponds to the naturalistic course of a group of disorders that is generally characterized by an episodic course. Furthermore, a decrease in distress among the study population during the observation period might be partially linked to the gradual suspension of lockdown measures during spring/summer 2020 and spring 2021 as a confounding factor. It has been shown that an escalation of lockdown measures during the COVID-19 pandemic and the rapid spreading of the novel coronavirus reliably lead to a deterioration of mental health in the general population within different cultural backgrounds around the world(30– 33). However, more recent studies hint at a persistence of distress and burden of psychopathology independent from periodic intensifications of lockdown measures^6^. In our study cohort, there was a marked difference in the decrease of symptom load between participants with episodic, mostly depressive disorders and chronic disorders like ADHD, strengthening the hypothesis that a significant part of the psychopathological improvement experienced by a distinct group of participants in our study truly represents a treatment effect.

Lastly, due to the naturalistic design of our study, there were neither a healthy control group nor placebo or sham treatments included. The latter might in any case have raised serious ethical concerns during a global pandemic. Nevertheless, we speculate that the participants in our study were generally more affected by psychological distress compared to the general population since the severity of their symptomatology crossed the threshold for diagnosing one of the above mentioned disorders and was eligible for psychiatric treatment within the German insurance system. However, we do not know the precise pre-pandemic level of psychopathology at baseline. Since a majority of patients that enrolled in our study had received psychiatric treatment at some time, however not immediately before the beginning of the study, they might have been at a greater risk for developing clinically relevant psychopathology during the COVID-19 pandemic, a finding that is also supported by results from a French cohort focusing on individuals with a history of depressive episodes(34).

## 5 Conclusions

Psychiatric telemedical treatment is an effective treatment option for patients with depressive disorders that yielded overall favorable outcomes in the observed group of patients. Although individuals diagnosed with depression in our sample had a higher load of psychopathology in the beginning, they profited most from telemedical treatment compared to participants with chronic neurodevelopmental disorders like ADHD who experienced an additional decline of well-being. The latter finding indicates that future research needs to concentrate on improving telemedical treatment options suited for chronic psychiatric conditions. Our study demonstrated a good match between patient-reported standardized measures of psychopathology and clinicians’ assessments during telemedical treatment, indicating that telemedical consultations could be a simple, economic and cost-effective but nevertheless reliable way of monitoring symptom severity and directing treatment choices during the treatment of depressive disorders. This is complemented by the overall high satisfaction of participants with the telemedical treatment they received. Therefore, current research on ecological momentary assessment (EMA) seems to open a promising new avenue towards personalized psychiatric telemedical treatment in the near future.

## Data Availability

All data are presented in the paper and the supplementary material. For details of the analysis with SPSS data can be made available on reasonable request.

## 7 Author Contributions

ASW and PP designed the study. TR, PP and ASW acquired data. AB developed the online survey tool. TR, TP and ASW analyzed data. OH, HT and AML advised and discussed data. PP, TP and ASW wrote the manuscript with the help of all authors. All authors have read and approved the manuscript.

## 9 Acknowledgments

The authors thank the service and telephone team of the outpatient clinic at the Central Institute of Mental Health Mannheim, Germany, under the lead of Volker Nitschke for their help with the recruitment of study subjects as well as Gerhard Kühne for the help with the analysis of data from the electronic medical records.

