## Supplementary Material for "Patients with affective disorders profit most from telemedical treatment: Evidence from a naturalistic patient cohort during the COVID-19 pandemic"

For

^†^shared first authorship

**Supplementary Table 1.** Table depicting psychopathological features which were reported in the medical record of the participants at the start of the telemedical treatment. Most participants revealed symptoms typical for affective disorders. N= number of subjects where information was found in the medical record. The percentage was calculated as (N/N responded)*100.

**Supplementary Table 2.** Detailed results of the two surveys of the SCL90-R before and 4-6 weeks after the beginning of the psychiatric treatment via telemedicine: The table provides the results of the three major indices of distress (GSI, PDSI and PST) as well as the subscales of the 9 psychopathological features including the number (N) of participants in the 1st and 2nd inquiry, the number of subjects with a t-values above 60 for the respective subcategory and their percentage relative to all participants ((N/N responded)*100). We also compared means between inquiry 1 and 2 for all subcategories and found a significant overall improvement (results inquiry 1 versus 2, ***P<0.001, Wilcoxon-Test was used).

**Supplementary Table 1**

|  | **N** | **abnormal** |
| --- | --- | --- |
| **Signs of Psychopathology** |  |  |
| **Vigilance** | **110** | **5(4,5 %)** |
| **Orientation** | **111** | **5(4,5 %)** |
| **Memory** | **109** | **29(26,6 %)** |
| **Perception** | **108** | **5(4,6 %)** |
| **Attention** | **108** | **56(51,9%)** |
| **Concentration** | **108** | **73(67,6)** |
| **Thought process** | **106** | **76(81,7)** |
| **Thought content** | **105** | **7(6,7 %)** |
| **Tricks of the senses** | **100** | **2(2,0 %)** |
| **Self-disorder (N=132)** | **99** | **8(8,1%)** |
| **Changes in mood** | **106** | **89(84,0 %)** |
| **Ability to experience joy** | **102** | **54(52,9 %)** |
| **Lack of drive** | **102** | **58(56,9 %)** |
| **Worries, Anxiety or Fear** | **101** | **65(64,4 %)** |
| **Intrusions** | **93** | **9(9,7 %)** |
| **Compulsive behavior** | **93** | **9(9,7 %)** |
| **Psychomotor function** | **96** | **29(30,2 %)** |
| **Changes in eating habit** | **85** | **17(20,0 %)** |
| **Sleep** | **98** | **68(69,4 %)** |
| **Libido** | **70** | **29(41,4 %)** |
| **Social interaction** | **69** | **9(13,0 %)** |
| **Self-harming behavior** | **132** | **9(6,8 %)** |
| **Illness insight** | **75** | **0(0,0 %)** |

**Supplementary Table 2**

|  | **1st inquiry (N)** | **T- Value ≥ 60** | **Mean** | **1st & 2nd inquiry (N)** | **1st** | **1st Mean** | **2nd T- Value ≥ 60 (N/%)** | **2nd Mean** |
| --- | --- | --- | --- | --- | --- | --- | --- | --- |
|  |  | **(N/%)** | **(SD)** |  | **T- Value ≥ 60 (N/%)** | **(SD)** |  | **(SD)** |
| **GSI** | 234 | 180 (77,0%) | 66,61 | 83 | 64 | 66.9 | 47 | 60,42*** |
|  |  |  | -10,31 |  | -77,10% | (8.45) | -56,60% | (10.02) |
| **PST** | 233 | 151 | 62,53 | 83 | 56 | 62,02 | 42 (50,6%) | 60,18** |
|  |  | -64,80% | -9,94 |  | -67,50% | -8,48 |  | -9,25 |
| **PSDI** | 231 | 175 | 65,29 | 83 | 63 | 65,12 | 42 | 59,45*** |
|  |  | -75,70% | -8,9 |  | -75,90% | -8,23 | -50,60% | -10,67 |
| **Somatization** | 226 | 125 (55,3%) | 60,28 | 81 | 46 | 61,17 | 35 (43,2%) | 56,78*** |
|  |  |  | -10,71 |  | -56,80% | -9,33 |  | -10,08 |
| **Obsessive-Compulsive** | 225 | 184 (81,8%) | 67,55 | 81 | 69 | 68,79 | 59 (72,8%) | 64,2*** |
|  |  |  | -10,77 |  | -85,20% | -9,52 |  | -10,08 |
| **Interpersonal Sensitivity** | 224 | 151 (67,4%) | 63,67 | 81 | 55 | 63,67 | 38 (46,9%) | 59,9*** |
|  |  |  | -11,33 |  | -67,90% | -9,81 |  | -11,11 |
| **Depression** | 195 | 155 (79,5%) | 67,08 | 83 | 70 | 67,75 | 48 (57,8%) | 63,37*** |
|  |  |  | -10,61 |  | -84,30% | -9,65 |  | -10,58 |
| **Anxiety** | 226 | 160 (70,8%) | 64,35 | 79 | 54 | 64,13 | 36 (45,6%) | 58,92*** |
|  |  |  | -10,95 |  | -68,40% | -9,38 |  | -10,22 |
| **Hostility** | 224 | 144 (64,3%) | 62,46 | 81 | 36 | 62,07 | 38 (42%) | 57,47*** |
|  |  |  | -10,75 |  | -44,40% | -10,89 |  | -10,12 |
| **Phobic Anxiety** | 227 | 134 (59,0%) | 61,32 | 82 | 50 | 60,67 | 34 (41,5%) | 56,89*** |
|  |  |  | -11,56 |  | -61,00% | -9,87 |  | -11,03 |
| **Paranoid Ideation** | 224 | 125 (55,8%) | 60,33 | 80 | 39 | 60,01 | 26 (32,5%) | 54,94*** |
|  |  |  | -11,38 |  | -48,80% | -9,79 |  | -10,48 |
| **Psychoticism** | 227 | 149 (65,6%) | 62,35 | 82 | 56 | 62,55 | 36 | 57,74*** |
|  |  |  | -9,99 |  | -68,30% | -8,31 | -43,90% | -10,05 |
